# Causal Relationships Between Soluble ST2, Heart Failure, and Sepsis: Bidirectional and Multivariable Mendelian Randomization Analyses

**DOI:** 10.64898/2026.01.20.26344488

**Authors:** Dongpo Liu, Yanxiang Sun

## Abstract

**BACKGROUND:** Soluble ST2 (sST2) predicts poor outcomes in heart failure (HF) and sepsis, but does it actually drive these conditions or just tag along for the ride? We used bidirectional Mendelian randomization (MR) to find out, testing six directional causal pathways among sST2, HF, and sepsis.

**METHODS AND RESULTS:** Our approach involved bidirectional two-sample Mendelian Randomization(MR) analyses drawing on genome-wide association study (GWAS) summary statistics: sST2 data came from deCODE Genetics (n=30,931), HF data from the Heart Failure Molecular Epidemiology for Therapeutic Targets(HERMES) Consortium (n=977,323), and sepsis data from FinnGen R12 (n=500,348) . We looked at six directions. IVW was the main method, with (Mendelian Randomization–Egger regression)MR-Egger, weighted median, (Mendelian Randomization Pleiotropy RESidual Sum and Outlier)MR-PRESSO sensitivity tests, and multivariable MR (MVMR). We did a cis-SNP analysis too, using only IL1RL1 variants. None of the six pathways showed a causal effect. Genetically predicted sST2 had no link to sepsis (OR: 1.01; 95% CI: 0.94–1.08; P=0.869) or HF (OR: 0.99; 95% CI: 0.92–1.07; P=0.867).cis-SNP analysis gave the same answer for sST2→sepsis (OR: 1.03; 95% CI: 0.98–1.09; P=0.223). MVMR adjusting for HF: still nothing (OR: 1.01; 95% CI: 0.93–1.09; P=0.862). HF and sepsis did not cause each other either - HF→sepsis (OR: 0.98; 95% CI: 0.79–1.23; P=0.882), sepsis→HF (OR: 1.07; 95% CI: 0.96–1.20; P=0.236). Going the other way, genetic risk for HF or sepsis did not affect sST2 levels.

**CONCLUSIONS:** We found no evidence that sST2 causes HF or sepsis. The picture that emerges is one where sST2 goes up because patients are sick-not the other way around. This makes sST2 a useful prognostic signal but probably not something worth targeting with drugs.We also found no direct causal link between HF and sepsis.

**Clinical Perspective:** *What Is New?:* This bidirectional Mendelian randomization study is the first to comprehensively examine potential causal relationships among soluble ST2, heart failure, and sepsis using genetic instruments. We found no evidence that genetically predicted sST2 levels causally influence the risk of heart failure or sepsis, despite strong observational associations reported in clinical studies. Similarly, genetic liability to heart failure or sepsis does not appear to causally affect circulating sST2 levels, suggesting sST2 elevation is a consequence rather than a cause of these conditions.

*What Are the Clinical Implications?:* These findings suggest that sST2 functions primarily as a prognostic biomarker reflecting disease severity rather than as a driver of pathophysiology, which has implications for its use in clinical decision-making. The strong observational associations between sST2 and adverse cardiovascular outcomes likely reflect confounding or reverse causation rather than direct causal effects. Drug development efforts targeting the ST2/IL-33 signaling pathway should consider that modulating sST2 levels may not directly prevent heart failure or improve sepsis outcomes.

---

Belonging to the interleukin-1 receptor family, soluble ST2 (sST2) has emerged as an important biomarker within cardiovascular medicine over recent years. Higher circulating sST2 concentrations show a consistent and strong relationship with poor outcomes among patients experiencing either acute or chronic heart failure (HF), which has prompted its adoption into clinical guidelines that address risk evaluation.^1,2^ Beyond its recognized utility in HF, researchers have noted sST2 elevation across various inflammatory states, sepsis being a prime example—here, sST2 levels track with both mortality rates and how severe the illness becomes.^3,4^ Why might this connection exist? The biological rationale appears solid: IL-33, the ligand that binds sST2, functions as a cellular ’alarmin’ that gets released when cells undergo stress or tissues sustain damage-phenomena lying at the heart of both HF and sepsis pathophysiology. When sST2 circulates in its soluble form, it essentially works as a decoy receptor. By trapping IL-33, it blocks the protective and anti-inflammatory signals that would otherwise flow through the membrane-bound ST2L receptor. Many researchers believe this interference with IL-33/ST2L signaling represents a crucial mechanism pushing disease forward.

Yet observational findings alone cannot prove causality, however compelling the prognostic data for sST2 might be. Confounding and reverse causation pose constant threats to such associations.One plausible scenario is that the high sST2 levels documented in HF and sepsis simply result from underlying pathophysiology-think myocardial stress, widespread inflammation, or endothelial damage-rather than driving these conditions forward.Consider that pro-inflammatory cytokines including TNF-α and IL-1β, both abundant during HF and sepsis, can boost sST2 production. Getting this distinction right matters tremendously: it shapes our understanding of what sST2 actually does biologically and whether targeting it therapeutically makes sense. Should sST2 genuinely cause disease, lowering its levels might help patients. Should it merely mark disease severity, then sST2 retains prognostic value but represents a poor drug target.

Mendelian randomization (MR) offers a powerful method for drawing causal conclusions by treating genetic variants as instrumental variables (IVs) that proxy for exposures of interest, free from typical confounding.⁵ Genetic variants get assigned at conception through random meiotic assortment, rendering them largely independent from environmental exposures and lifestyle choices that muddy observational research. When applied to large-scale GWAS summary data, MR generates evidence regarding whether a putative risk factor truly causes a disease outcome—assuming the method’s underlying requirements hold.

For this investigation, we carried out a thorough bidirectional two-sample MR study probing the causal web connecting circulating sST2, HF, and sepsis. Two main questions drove our work: Does genetically elevated sST2 actually increase someone’s likelihood of developing HF or sepsis? And running the analysis backward, does genetic susceptibility to HF or sepsis alter circulating sST2 concentrations?Through examining causation in both directions, we aimed to pin down whether sST2 functions as a genuine causal agent or simply a passive marker within the clinical overlap between HF and sepsis.

## METHODS

### Overall Study Design

This study used bidirectional two-sample MR to look at causal links between sST2, heart failure, and sepsis. MR uses genetic variants as proxies for the exposure - the idea being that genes are assigned randomly at conception, so they sidestep the confounding problems you get with regular observational studies. We tested all six possible directions: sST2→sepsis, sST2→HF, HF→sepsis, sepsis→HF, HF→ sST2, and sepsis→sST2. For MR to work, the genetic instruments need to actually associate with the exposure (relevance), not be linked to confounders (independence), and only affect the outcome through the exposure (exclusion restriction).We used publicly available GWAS summary statistics and followed STROBE-MR guidelines.⁶

### Data Sources and Genetic Instrument Selection

We gathered summary statistics for genetic variants showing robust trait associations from leading GWAS consortia and databases (Table 1 provides details). We picked datasets with minimal participant overlap to avoid winner’s curse problems (Table S1). The sST2 data came from Iceland, sepsis from Finland—no shared participants there. The HERMES consortium drew on multiple European cohorts, but Icelandic and Finnish contributions were negligible, keeping overlap risk low for those analyses too. Our SNP selection followed standard genome-wide significance (P < 5×10⁻⁸) for most exposures. For sepsis, the standard threshold left us with too few SNPs to work with, so we relaxed it to P < 5×10⁻⁶. This is not unusual when dealing with smaller GWAS—others have done the same.^11^ The tradeoff is that looser thresholds can let weaker instruments slip through, which would bias results toward the null. We took several steps to guard against this: the mean F-statistic for our sepsis instruments came in at 24.0, well clear of the conventional cutoff of 10, and we leaned on sensitivity methods (MR-Egger, weighted median, MR-PRESSO) that hold up better when some instruments are shaky. Still, readers should keep this caveat in mind when interpreting sepsis-related findings. We then applied linkage disequilibrium clumping (r² < 0.001 within 10,000 kb windows) referenced against 1000 Genomes European data to guarantee independent SNPs. Any palindromic SNPs presenting ambiguous allele frequencies got dropped.

**Table 1.** Description of Data Sources for Mendelian Randomization Analysis

| Phenotype | Data Source | Sample Size | Population | Reference |
| --- | --- | --- | --- | --- |
| sST2 (Soluble ST2) | deCODE Genetics | 30,931 | European (Icelandic) | Sun et al. 2018 |
| Heart Failure | HERMES Consortium | 977,323 (47,309 cases) | European (Multi-cohort) | Shah et al. 2020 |
| Sepsis | FinnGen R12 | 500,348 (21,547 cases) | Finnish | FinnGen 2023 |

Genetic instruments for circulating sST2 levels were derived from a GWAS of 30,931 individuals of Icelandic ancestry conducted by deCODE Genetics.⁷ Summary statistics for HF were obtained from the Heart Failure Molecular Epidemiology for Therapeutic Targets (HERMES) consortium, a major international effort focused on the genetics of HF in individuals of European descent, which included 47,309 cases and 930,014 controls.⁸ Genetic associations for sepsis were sourced from FinnGen Release 12, which included 21,547 cases and 478,801 controls of Finnish ancestry.⁹

We gauged IV strength via F-statistics, computed as F = (beta/se) ² . Values exceeding 10 conventionally signal sufficient instrument strength, guarding against weak instrument problems. Our analyses consistently showed mean F-statistics well above this cutoff, validating the instruments for MR purposes.

### Statistical Analysis

We ran all statistical procedures through R (version 4.1.0), relying primarily on the TwoSampleMR and MRPRESSO packages.

Our main analysis used random-effects IVW. Basically, you take each SNP’s ratio of outcome-to-exposure effect and combine them. Works well when the instruments are all valid, or when pleiotropy at least balances out.

Robustness checking and formal assumption testing required several sensitivity analyses. MR-Egger regression yields causal estimates that remain valid even under directional pleiotropy; its intercept term signals average pleiotropic effects across instruments, with P < 0.05 flagging problematic directional pleiotropy that would breach the exclusion restriction. Weighted median estimation stays consistent provided less than half the statistical weight comes from invalid instruments.MR-PRESSO tackles pleiotropy head-on by spotting and excluding outlier SNPs.

We ran an additional cis-SNP analysis for the sST2 exposure, restricting attention to cis-acting variants lying within ±500kb of IL1RL1 (chr2: 102,000,000-103,500,000 bp)—the gene encoding sST2. Using only these local variants limits the analysis to SNPs with clear biological mechanisms for affecting sST2, thereby shrinking horizontal pleiotropy risks.

We additionally performed multivariable Mendelian randomization (MVMR) to estimate the direct causal effect of sST2 on sepsis while accounting for potential mediation or confounding through heart failure. MVMR extends standard two-sample MR by including genetic instruments for multiple exposures simultaneously, allowing estimation of the direct effect of each exposure independent of the others. We used instruments for both sST2 and HF, harmonized with the sepsis outcome, and applied the MVMR-IVW method implemented in the MVMR package. Conditional F-statistics were calculated to assess instrument strength for each exposure after accounting for the other.

We used Cochran’s Q to check if the SNP estimates were all pointing in the same direction. If Q is significant (P < 0.05), something might be off - maybe pleiotropy, maybe something else. For binary outcomes like HF and sepsis, we report odds ratios with 95% CIs. For sST2 (continuous), we report betas - basically how much sST2 changes in standard deviations for each log-odds increase in disease risk.

## RESULTS

### Genetic Instrument Strength and Validity

After filtering, we retained 9 independent SNPs for sST2, 12 for heart failure, and 13 for sepsis. Dataset harmonization left 7 to 10 SNPs available for each specific MR analysis. Instrument strength looked excellent: mean F-statistics reached 329.2 for sST2, 40.8 for HF, and 24.0 for sepsis—all comfortably surpassing the threshold of 10. Even the lowest individual SNP F-statistic exceeded 10, reinforcing confidence in our instruments.

### Bidirectional Mendelian Randomization Analyses

Table 2 compiles the IVW findings for all six pathways, with Figure 2 presenting them graphically.

**Figure 1.**
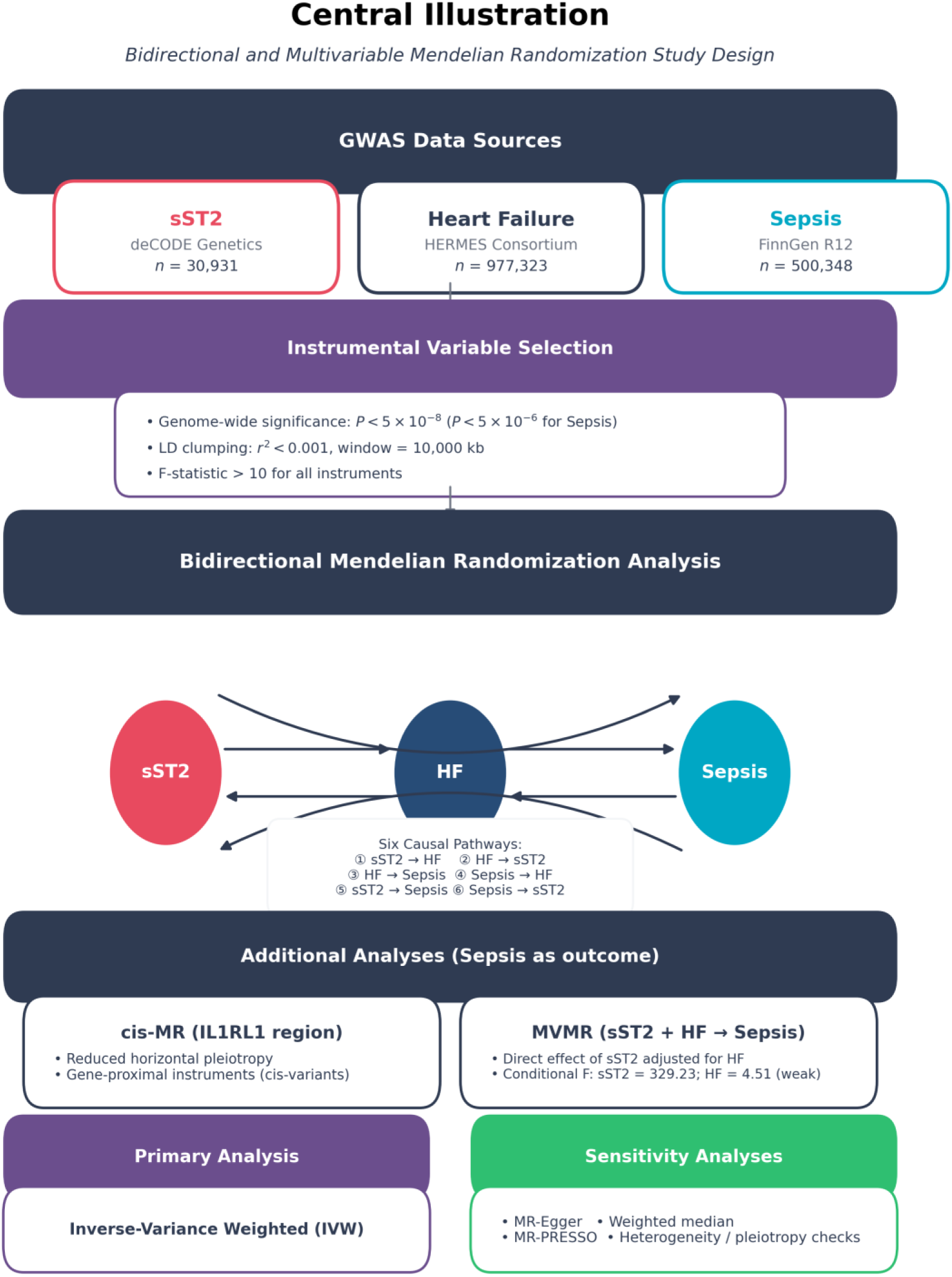
Central Illustration of the Study Design. The study leveraged summary-level data from three large-scale GWAS for sST2, heart failure, and sepsis. After selecting robust genetic instruments, a bidirectional Mendelian randomization analysis was conducted to evaluate six potential causal pathways. The primary analysis used the IVW method, with a suite of sensitivity analyses to test for pleiotropy.

**Figure 2.**
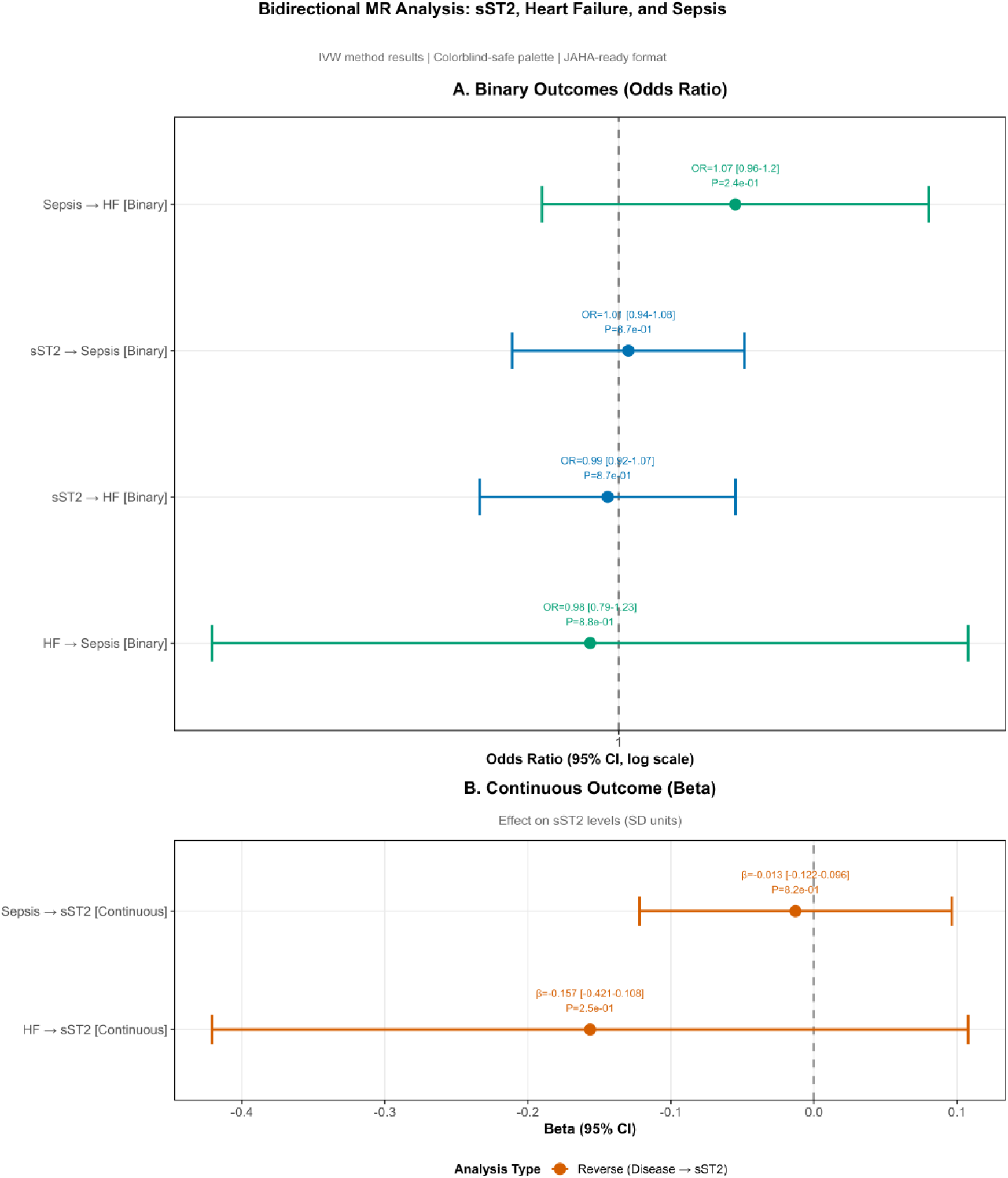
Forest plot summarizing the bidirectional Mendelian randomization results. Panel A covers binary outcomes—whether sST2, HF, or sepsis affects the others—shown as odds ratios from the IVW method. Panel B covers continuous outcomes, showing how genetic liability to HF or sepsis relates to sST2 blood levels (beta = SD change per log-odds increase in disease risk). Error bars are 95% CIs. The dashed line marks the null. None of the six pathways hit significance.

**Figure 3.**
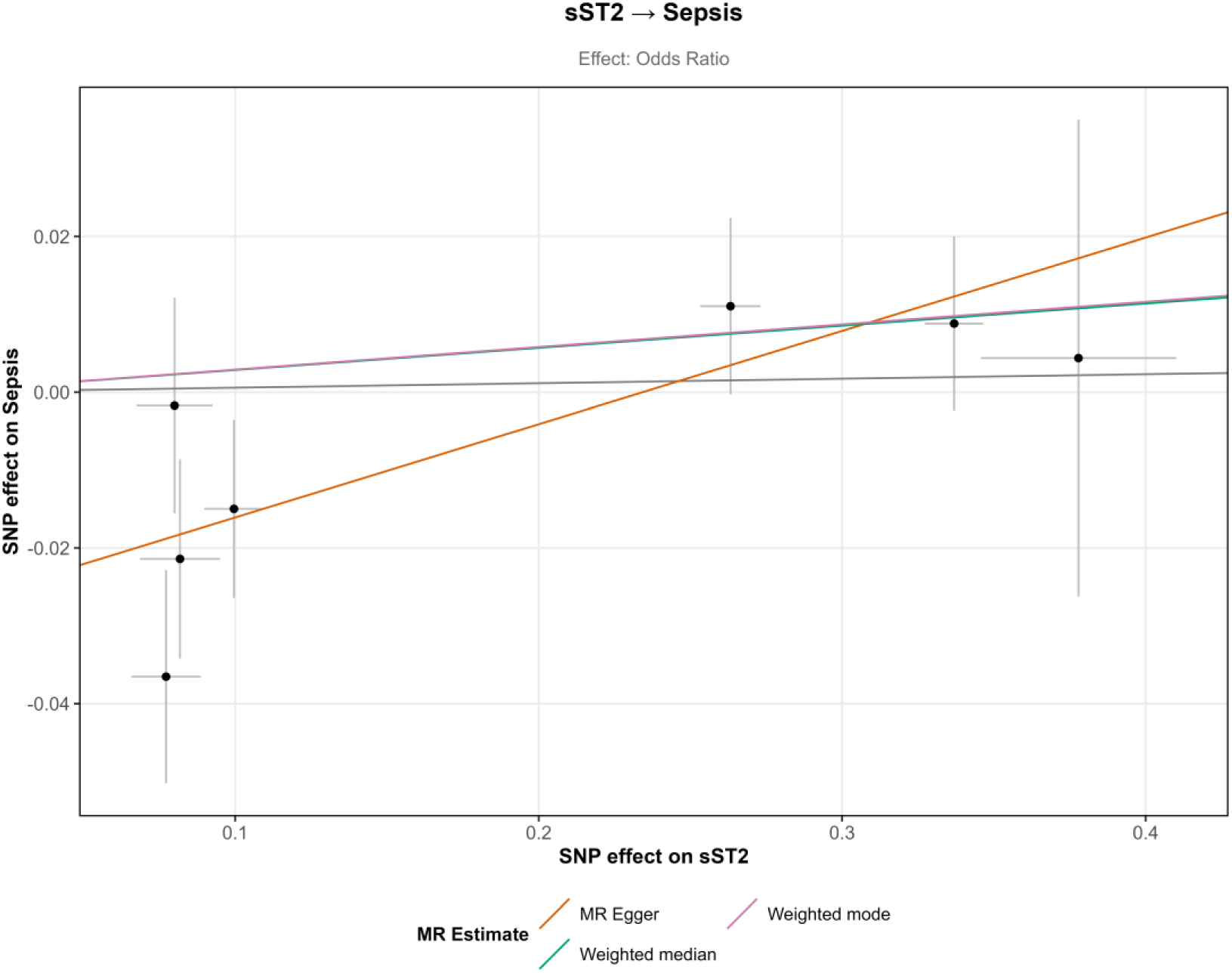
Scatter plot for the causal effect of sST2 on sepsis. Each point represents a single SNP, with the x-axis showing the SNP effect on sST2 and the y-axis showing the SNP effect on sepsis. The slopes of the lines represent the causal estimates from different MR methods. The SNP estimates scatter around zero with no clear trend—exactly what you would expect if there is no causal effect.

**Figure 4.**
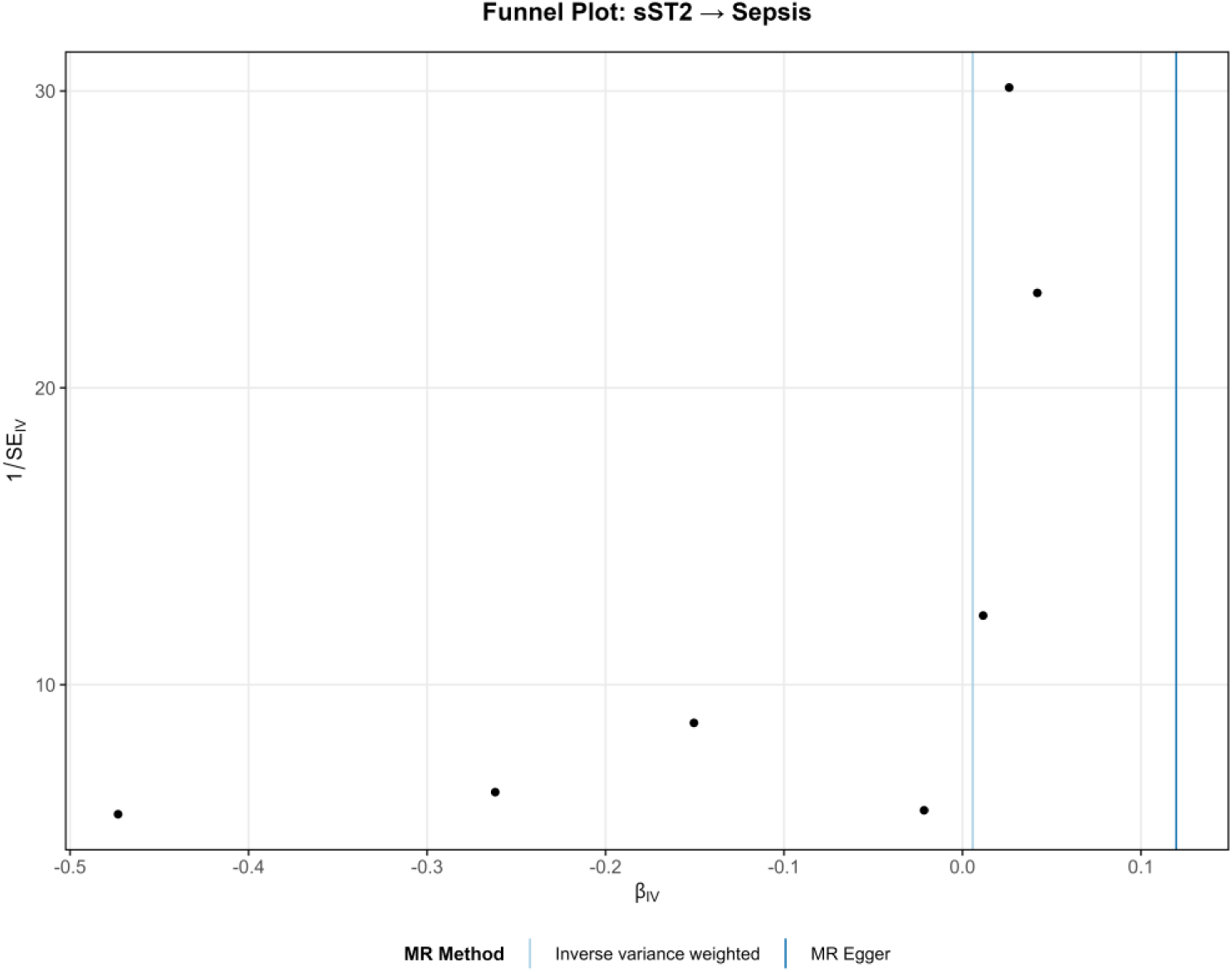
Funnel plot for the causal effect of sST2 on sepsis. The funnel plot displays the individual SNP causal estimates (x-axis) against their precision (1/SE, y-axis). Asymmetry in the funnel plot may indicate the presence of directional pleiotropy. The plot appears reasonably symmetric around the IVW estimate, providing visual reassurance against major directional pleiotropy.

**Figure 5.**
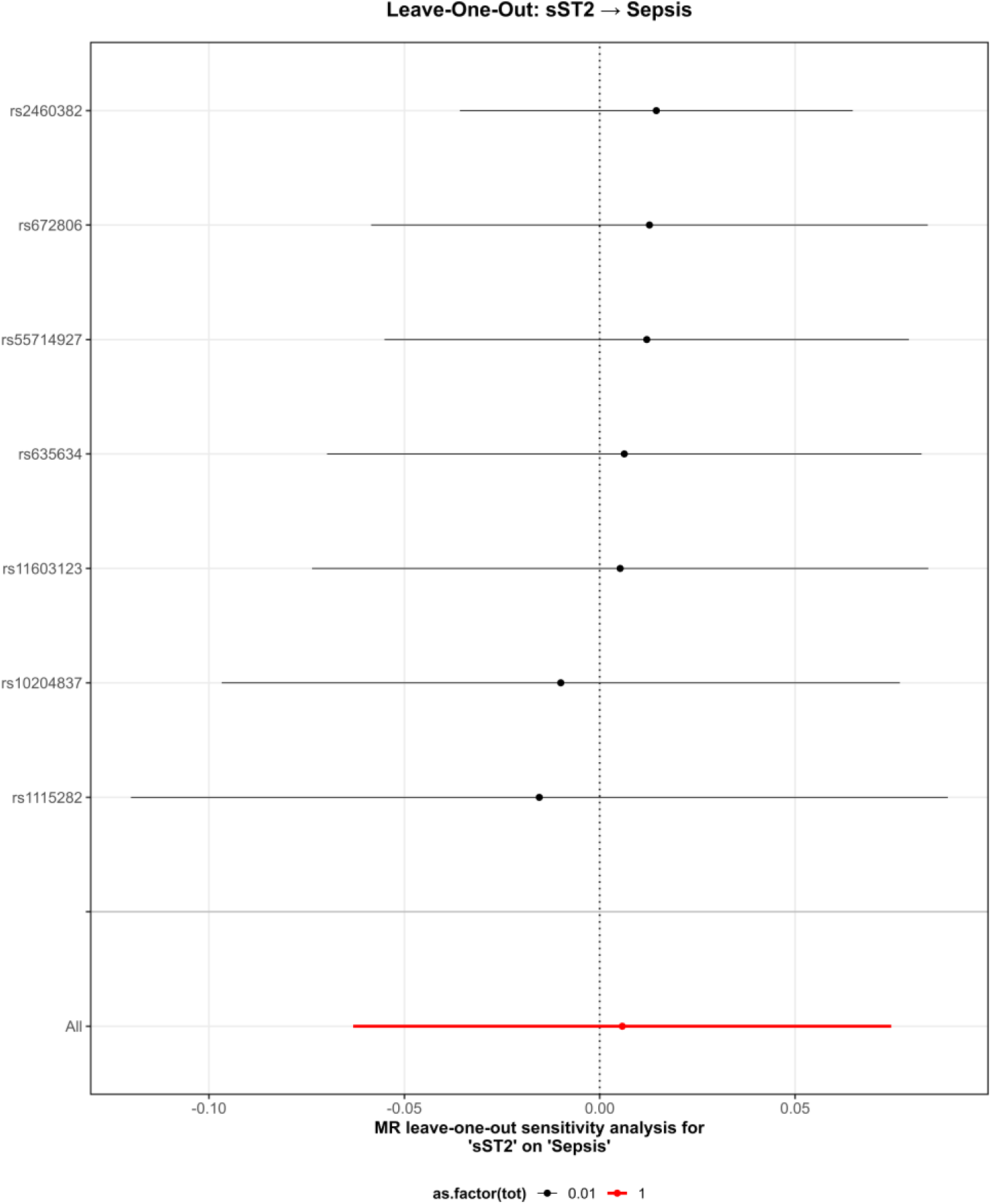
Leave-one-out sensitivity analysis for the causal effect of sST2 on sepsis. Each row shows the IVW estimate when the corresponding SNP is excluded from the analysis. The red line at the bottom shows the overall estimate using all SNPs. The overall estimate barely budges when any single SNP is dropped, so no single variant is driving the null result.

**Table 2.**
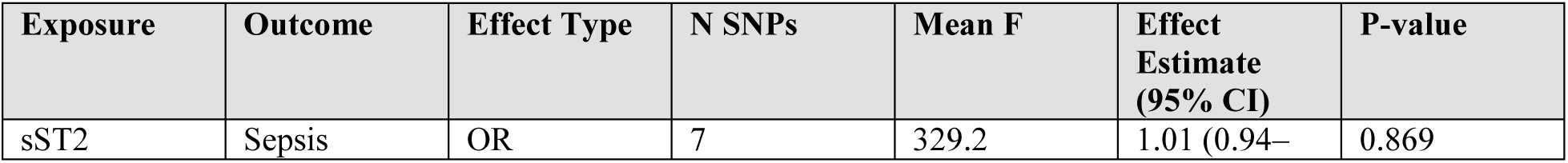
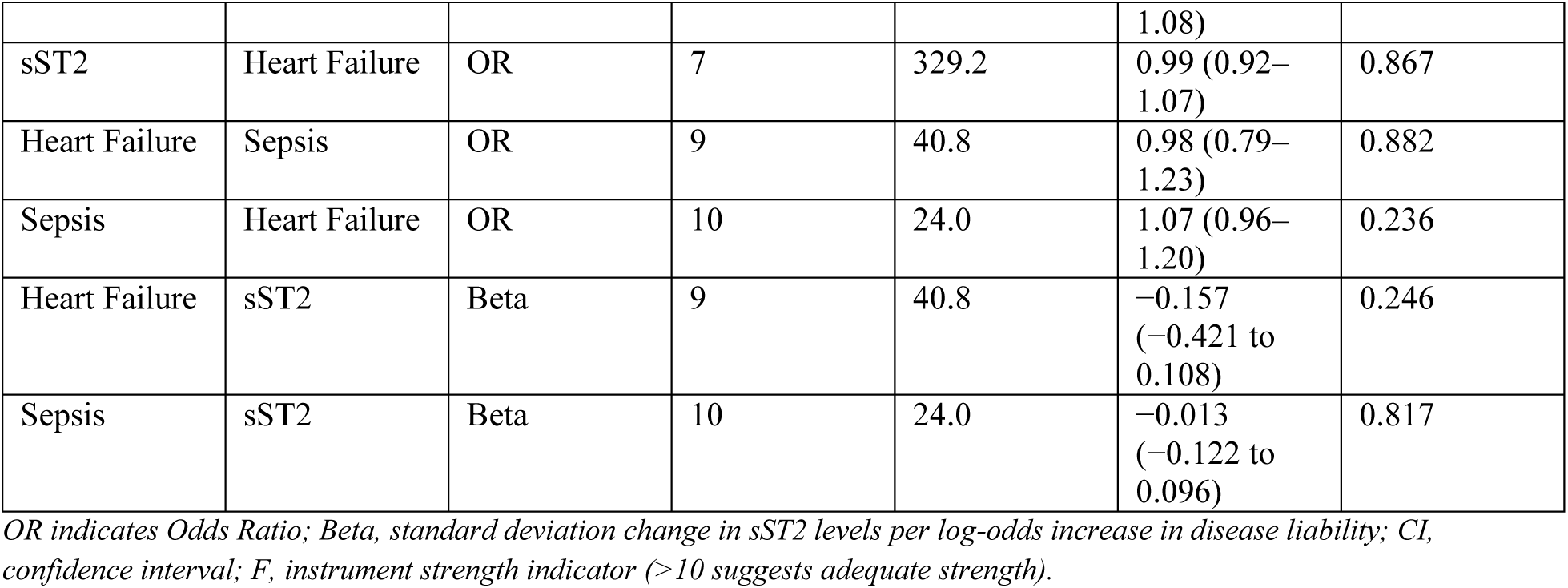
Primary Mendelian Randomization Results Using the Inverse-Variance Weighted Method

#### Causal Effect of sST2 on Heart Failure and Sepsis

The main IVW analysis turned up no causal connection between genetically instrumented sST2 and either sepsis (OR: 1.01; 95% CI: 0.94–1.08; P=0.869) or heart failure (OR: 0.99; 95% CI: 0.92–1.07; P=0.867). Every sensitivity method—MR-Egger, weighted median, MR-PRESSO—yielded consistent null findings, bolstering result robustness. Point estimates clustered near 1 across all approaches, and tight confidence intervals indicated decent statistical power for detecting any clinically relevant effect had one existed.

#### cis-SNP Sensitivity Analysis

For added validation addressing pleiotropy concerns, we conducted a cis-SNP analysis using just two SNPs (rs10204837, rs1115282) mapping to the IL1RL1 region on chromosome 2. These cis variants directly influence sST2 through biologically obvious mechanisms and showed outstanding instrument strength (mean F: 969.2). Results for the sST2-to-sepsis pathway echoed the primary analysis—no causal signal (OR: 1.03; 95% CI: 0.98–1.09; P=0.223)—offering compelling evidence that sST2 does not drive sepsis (Table 3).

**Table 3.** Results of cis-SNP Mendelian Randomization Analysis

| Analysis | Region | N SNPs | Method | Beta | SE | OR (95% CI) | P-value |
| --- | --- | --- | --- | --- | --- | --- | --- |
| sST2 → Sepsis | IL1RL1 (chr2) | 2 | IVW | 0.032 | 0.026 | 1.03 (0.98–1.09) | 0.223 |

*cis-SNPs were defined as variants within ±500kb of the IL1RL1 gene (chr2: 102,000,000-103,500,000 bp). Mean F-statistic for cis-SNPs: 969.2.*

#### Multivariable Mendelian Randomization Analysis

To assess whether the effect of sST2 on sepsis might be mediated or confounded by heart failure, we conducted multivariable Mendelian randomization (MVMR) including both sST2 and HF as exposures with sepsis as the outcome. The MVMR analysis used 7 SNPs that were valid instruments for both exposures after harmonization. The conditional F-statistic for sST2 remained strong (329.23), indicating adequate instrument strength even after accounting for HF. However, the conditional F-statistic for HF was low (4.51), suggesting potential weak instrument bias for this exposure in the MVMR framework. After adjusting for HF, sST2 showed no direct causal effect on sepsis (OR: 1.01; 95% CI: 0.93–1.09; P=0.862). Similarly, HF showed no direct effect on sepsis independent of sST2 (OR: 1.12; 95% CI: 0.53–2.38; P=0.768), though this estimate should be interpreted cautiously given the weak instrument strength. These MVMR results reinforce the univariable findings and suggest that the null association between sST2 and sepsis is not confounded by heart failure (Table 4).

**Table 4.** Multivariable Mendelian Randomization Results: Direct Effects on Sepsis

| Exposure | No. of SNPs | Conditional F | Beta (SE) | OR (95% CI) | P value |
| --- | --- | --- | --- | --- | --- |
| sST2 | 7 | 329.23 | 0.007 (0.038) | 1.01 (0.93–1.09) | 0.862 |
| Heart Failure | 7 | 4.51* | 0.113 (0.384) | 1.12 (0.53–2.38) | 0.768 |

*Outcome: Sepsis. Conditional F-statistics >10 indicate adequate instrument strength in MVMR context. \*Conditional F <10 suggests potential weak instrument bias; results should be interpreted with caution. OR, odds ratio; CI, confidence interval; SE, standard error.*

#### Bidirectional Causal Effects Between Heart Failure and Sepsis

No causal relationship emerged between HF and sepsis in either direction. HF genetic liability showed no association with sepsis risk (IVW OR: 0.98; 95% CI: 0.79–1.23; P=0.882). Going the other way, sepsis genetic liability did not causally affect HF risk (IVW OR: 1.07; 95% CI: 0.96–1.20; P=0.236). The sepsis-to-HF point estimate sat slightly above 1, true, but the confidence interval crossed the null and the P-value missed significance.

#### Causal Effects of Heart Failure and Sepsis on sST2 Levels

Running MR in reverse, we asked whether genetic risk for HF or sepsis might shape sST2 concentrations. The answer was negative. Genetic HF liability (Beta: -0.157; 95% CI: -0.421 to 0.108; P=0.246) and genetic sepsis liability (Beta: -0.013; 95% CI: -0.122 to 0.096; P=0.817) both showed null associations with circulating sST2.These results strengthen the interpretation that sST2 marks rather than causes these conditions.

### Sensitivity Analyses and Pleiotropy Assessment

Sensitivity estimates from MR-Egger, weighted median, and weighted mode largely mirrored the primary IVW results, lending additional support to the null findings (Table 5). Effect directions and magnitudes tracked closely across methods, with no approach yielding significance.

**Table 5.** Summary of Sensitivity Analyses for All Mendelian Randomization Pathways

| Analysis | Method | Beta (SE) | P-value |
| --- | --- | --- | --- |
| sST2 → Sepsis | IVW | 0.006 (0.035) | 0.869 |
|  | MR-Egger | 0.120 (0.044) | 0.043 |
|  | Weighted median | 0.029 (0.027) | 0.294 |
|  | Weighted mode | 0.029 (0.026) | 0.315 |
| sST2 → Heart Failure | IVW | -0.006 (0.039) | 0.867 |
|  | MR-Egger | 0.080 (0.066) | 0.281 |
|  | Weighted median | 0.006 (0.021) | 0.774 |
|  | Weighted mode | 0.011 (0.019) | 0.602 |
| Heart Failure → Sepsis | IVW | -0.017 (0.114) | 0.882 |
|  | MR-Egger | -0.185 (0.360) | 0.623 |
|  | Weighted median | 0.048 (0.111) | 0.664 |
|  | Weighted mode | 0.081 (0.133) | 0.561 |
| Sepsis → Heart Failure | IVW | 0.069 (0.058) | 0.236 |
|  | MR-Egger | 0.167 (0.133) | 0.244 |
|  | Weighted median | 0.122 (0.064) | 0.057 |
|  | Weighted mode | 0.154 (0.082) | 0.092 |

IVW indicates Inverse-Variance Weighted; SE, Standard Error.

Cochran’s Q flagged significant heterogeneity in a couple of analyses—sST2 on HF (Q=31.44, P < 0.001) and HF on sST2 (Q=36.74, P < 0.001). What might be going on? A few possibilities. Our sST2 instruments include both cis-variants near IL1RL1 and trans-variants scattered elsewhere in the genome; these may work through different biological routes and give different effect sizes. Population differences between datasets—Icelandic for sST2, pan-European for HF—could also contribute. And some instruments might be doing things on the side (pleiotropy). The reassuring part: MR-Egger intercepts stayed non-significant for most pathways (except sST2→sepsis at P=0.029), which means any pleiotropy is probably balanced rather than pushing results in one direction. MR-PRESSO flagged a handful of outlier SNPs, but dropping them left the null findings intact. The consistency of null results across IVW, MR-Egger, weighted median, and MR-PRESSO gives us confidence that heterogeneity is not driving our conclusions (Table 6).

**Table 6.** Assessment of Heterogeneity and Horizontal Pleiotropy

| Analysis | Cochran’s Q | Q df | Q P-value | Egger Intercept | Egger P-value | PRESSO Outliers |
| --- | --- | --- | --- | --- | --- | --- |
| sST2 → Sepsis | 13.18 | 6 | 0.040 | −0.028 | 0.029 | 0 |
| sST2 → Heart Failure | 31.44 | 6 | <0.001 | −0.022 | 0.185 | 1 |
| Heart Failure<br>→ Sepsis | 18.72 | 8 | 0.016 | 0.012 | 0.636 | 1 |
| Sepsis →<br>Heart Failure | 15.38 | 9 | 0.081 | −0.009 | 0.434 | 0 |
| Heart Failure<br>→ sST2 | 36.74 | 8 | <0.001 | −0.013 | 0.647 | 1 |
| Sepsis → sST2 | 7.52 | 9 | 0.583 | 0.018 | 0.155 | 0 |

Significant heterogeneity ( $Q$ $P$ -value < 0.05) was observed in several analyses. However, the MR-Egger intercept test for directional pleiotropy was non-significant ( $P$ > 0.05) for most analyses.

## DISCUSSION

Our bidirectional MR work yielded consistently negative results—sST2 showed no causal link to HF or sepsis in any direction we tested. The reverse analyses told a similar story: carrying genetic risk for HF or sepsis does not seem to push sST2 levels up or down. We also came up empty when looking for a causal connection between HF and sepsis directly. So what do we make of the well-known association between elevated sST2 and poor prognosis in HF and sepsis? Recent studies continue to confirm the strong prognostic value of sST2—Ye et al. demonstrated that sST2 achieved an AUC of 0.912 for predicting sepsis mortality, comparable to SOFA and APACHE II scores.¹² A 2025 comparative analysis by Davini et al. further showed that sST2 serves as a robust predictor of mortality (AUC 0.87) and hospital readmission in patients with overlapping acute heart failure and sepsis.¹³ Our findings challenge the notion that sST2 sits in the causal chain. Despite strong observational associations with adverse outcomes,⁷,⁸,¹²,¹³ our MR results suggest that elevated sST2 most likely reflects disease severity rather than driving disease progression. Why would sST2 go up in HF and sepsis if it is not causing them? The biology offers a straightforward answer. When tissues get stressed—cardiomyocytes stretched, endothelium inflamed—cells ramp up sST2 production. Pro-inflammatory cytokines like TNF-α and IL-1β, which run rampant in both HF and sepsis, directly stimulate sST2 release.¹⁴ So sST2 is less a villain and more a messenger reporting back on how bad things have gotten. Yes, sST2 mops up IL-33 and blunts some protective signaling through ST2L.⁹ But this “decoy receptor” activity might itself be a compensatory brake on runaway inflammation rather than a disease accelerant.¹⁵ Think of sST2 as a smoke detector: it tells you there is a fire, but smashing it will not put out the flames.

We gained extra confidence from the cis-SNP analysis. By zeroing in on variants sitting right next to IL1RL1—the gene behind sST2—we cut way down on pleiotropy concerns and got estimates with clearer biological meaning. The result? Still no hint of causality (OR: 1.03; 95% CI: 0.98–1.09; P=0.223). sST2 does not appear to drive sepsis.

Our MVMR analysis provided another layer of evidence by examining whether the sST2-sepsis relationship might be confounded or mediated by heart failure. Even after accounting for HF, sST2 showed no direct effect on sepsis (OR: 1.01; 95% CI: 0.93–1.09; P=0.862). The conditional F-statistic for sST2 remained robust at 329.23, confirming strong instrument validity in this multivariable framework. The HF estimate in MVMR (OR: 1.12; P=0.768) should be interpreted cautiously given its weak conditional F-statistic (4.51), but the key finding is that adjusting for this potential confounder did not unmask any hidden sST2 effect. This triangulation—univariable MR, cis-SNP analysis, and MVMR all pointing to zero—strengthens the case that sST2 truly lacks causal influence on sepsis.

The lack of a genetic causal link between HF and sepsis caught our attention too. Doctors see these two problems together all the time, and textbooks describe elaborate mechanistic crosstalk. Sepsis hammers the heart through cytokine storms, mitochondrial shutdown, and microvascular chaos; HF, for its part, leaves patients vulnerable to infections because their immune defenses are weakened.¹⁰ Interestingly, a recent MR study by Ren et al. (2024) also explored the HF-sepsis relationship using similar genetic approaches combined with MIMIC-IV observational data, finding that while HF increases sepsis-related mortality in clinical settings, the genetic causal evidence remains limited.¹⁶ Their network MR analysis identified nerve growth factor as a potential mediating pathway, highlighting the complexity of HF-sepsis interactions that may operate through acquired rather than genetic mechanisms. Yet when we look through the MR lens, genetic loading for one condition does not raise the odds of the other. What brings HF and sepsis together in the same patient may have more to do with shared risk factors—getting older, having diabetes or kidney trouble—or with one acute illness setting off another, rather than any deep genetic link.

### Clinical Implications

What should clinicians and researchers take away? Keep ordering sST2 when you want to know how much trouble a patient is in - it still works for risk stratification. A high sST2 still means trouble ahead. You can pair it with procalcitonin for sepsis¹⁷ or throw it into a panel with BNP and troponins for HF.¹⁸ But here is the key point: predicting bad outcomes is not the same as causing them. LDL is causal - lower it with statins and patients actually do better. sST2? Not so much, based on what we found. Think of it as a thermometer, not a thermostat. You would not treat a fever by smashing the thermometer. The IL-33/ST2 pathway is also messier than people thought - IL-33 seems to help clear bacteria early on but might also contribute to immunosuppression later.¹⁵ That makes it a tricky drug target. Pharma companies eyeing this pathway should probably look elsewhere, or at least be careful. And more generally, this is exactly why MR matters - it helps you figure out what is actually causal versus what just looks causal in observational data.

### Strengths and Limitations

On the plus side, running MR in both directions let us test causality without the usual worries about confounders or getting cause and effect backwards. We also tapped into some of the biggest genetic datasets out there, which kept our confidence intervals tight.And the fact that MR-Egger, MR-PRESSO, and cis-SNP analyses all pointed the same way gives us more confidence that the null results are real. Picking datasets with little participant overlap (Table S1) helped us dodge winner’s curse problems.

A few caveats deserve mention. We stuck to European-ancestry samples—Icelandic, Finnish, pan-European—to keep population structure from muddying the analysis. The flip side is that we cannot say whether these findings hold in East Asian, African, or other populations. Genetic architecture can differ, and so might causal relationships. The sepsis GWAS was the smallest of the three (about 21,500 cases versus 47,000 for HF). We also had to loosen the P-value threshold to get enough instruments. Both factors limit our power to detect small effects. If sST2 has a weak but real causal role in sepsis, we might have missed it. Sepsis heterogeneity presents another challenge—emerging research emphasizes that sepsis comprises multiple endotypes with distinct pathophysiological mechanisms, and single biomarkers or genetic approaches may not fully capture this complexity.¹⁹,²⁰ MR tells you what happens over a lifetime of genetically higher or lower exposure. It does not capture what a sudden spike in sST2 does over hours or days—say, during septic shock. Acute sST2 surges could have short-term effects that our lifetime genetic proxies simply cannot see. That question will need different study designs to answer. And while our sensitivity analyses did not scream pleiotropy, we cannot rule it out entirely. The MR-Egger intercept for sST2→sepsis did reach nominal significance (P=0.029), though the overall null held across methods.

### Conclusions

Bottom line: we found no genetic evidence that sST2 causes HF or sepsis, and no evidence that HF and sepsis cause each other. A focused cis-SNP analysis backed up these negative findings.Taken together, the data paint sST2 as a strong prognostic signal—a thermometer reading the fever, not the infection causing it.Going forward, researchers and clinicians can keep relying on sST2 to stratify risk, but the search for treatable causal mechanisms in HF and sepsis will need to look elsewhere.

## SOURCES OF FUNDING

No specific funding from public, commercial, or nonprofit agencies supported this work.

## DISCLOSURES

None.

## DATA AVAILABILITY

All genome-wide association study summary statistics used in this study are publicly available. The sST2 GWAS data from deCODE Genetics are available through the GWAS Catalog. Heart failure GWAS summary statistics from the HERMES consortium are accessible at the GWAS Catalog (accession GCST009541). Sepsis GWAS data are from FinnGen Release 12, available at https://r12.finngen.fi/. The analytical code used to perform Mendelian randomization analyses is available from the corresponding author upon reasonable request.

## Data Availability

All GWAS summary statistics used in this study are publicly available through the GWAS Catalog (deCODE Genetics, HERMES consortium) and the FinnGen R12 website. Analytical code is available from the corresponding author upon reasonable request.

## ACKNOWLEDGMENTS

We gratefully acknowledge the deCODE Genetics, HERMES consortium, and FinnGen investigators and participants who made their summary statistics publicly accessible.

---

**Figure S1.**
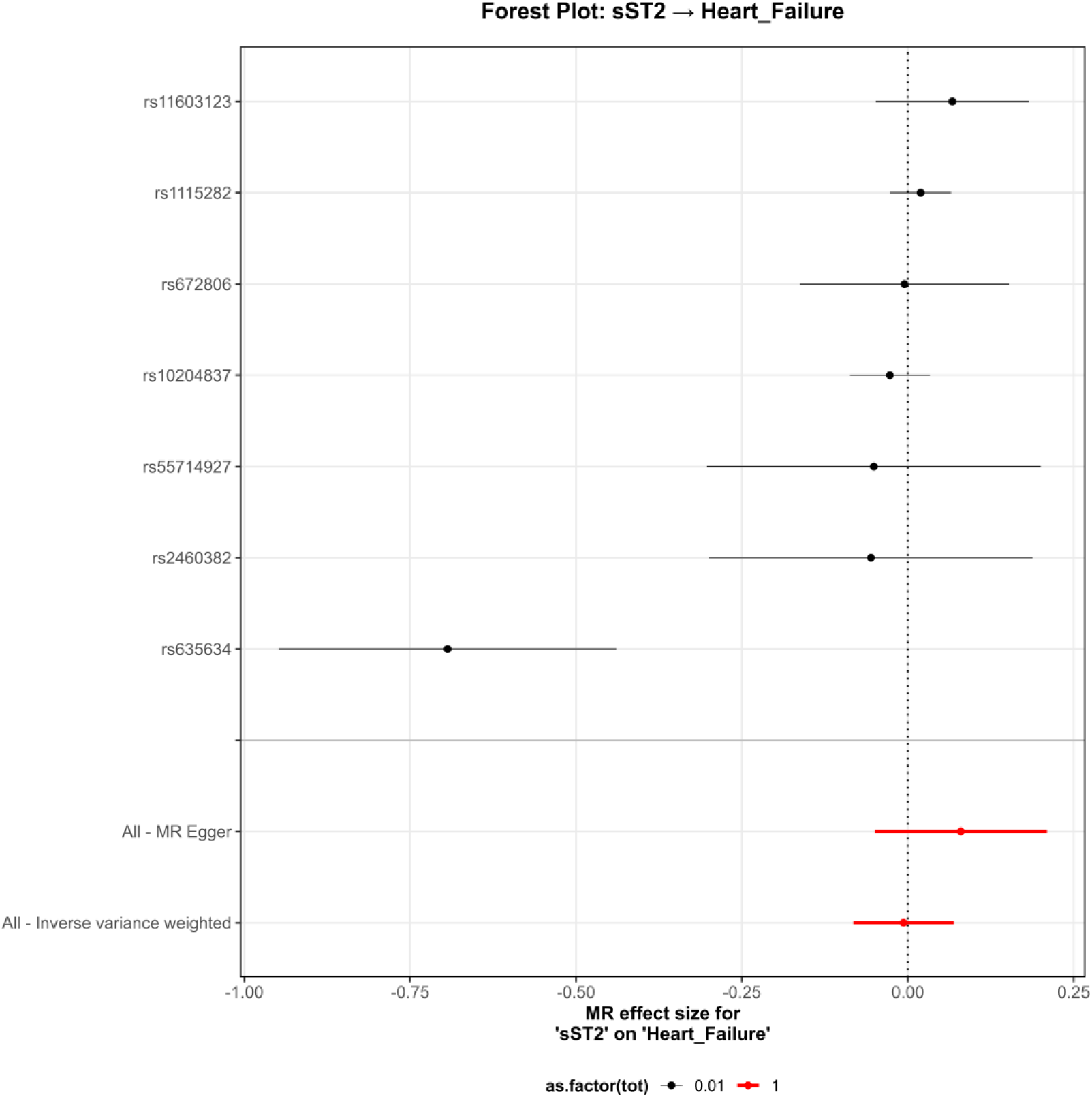
Forest plot for the causal effect of sST2 on heart failure.

**Figure S2.**
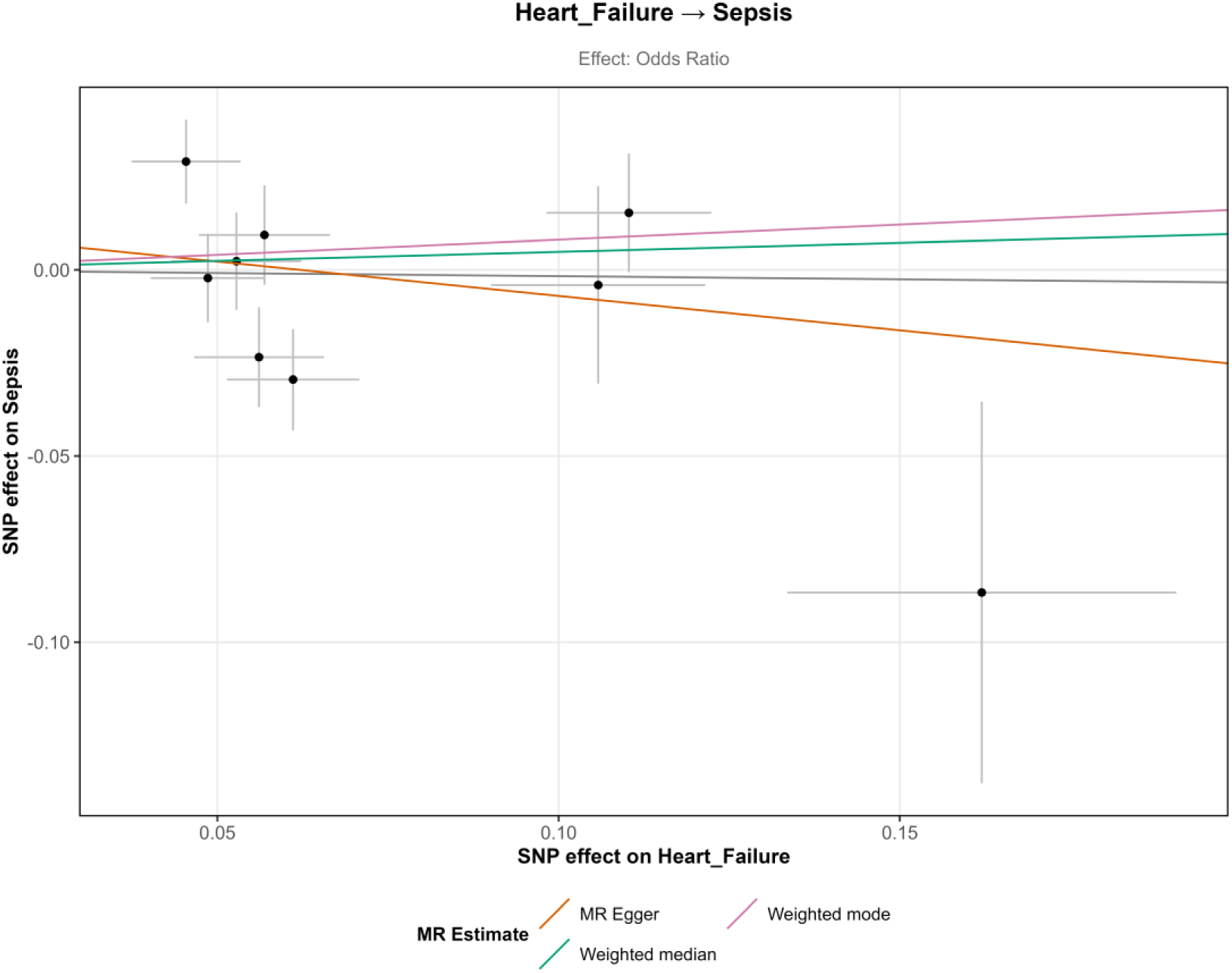
Scatter plot for the causal effect of heart failure on sepsis.

**Figure S3.**
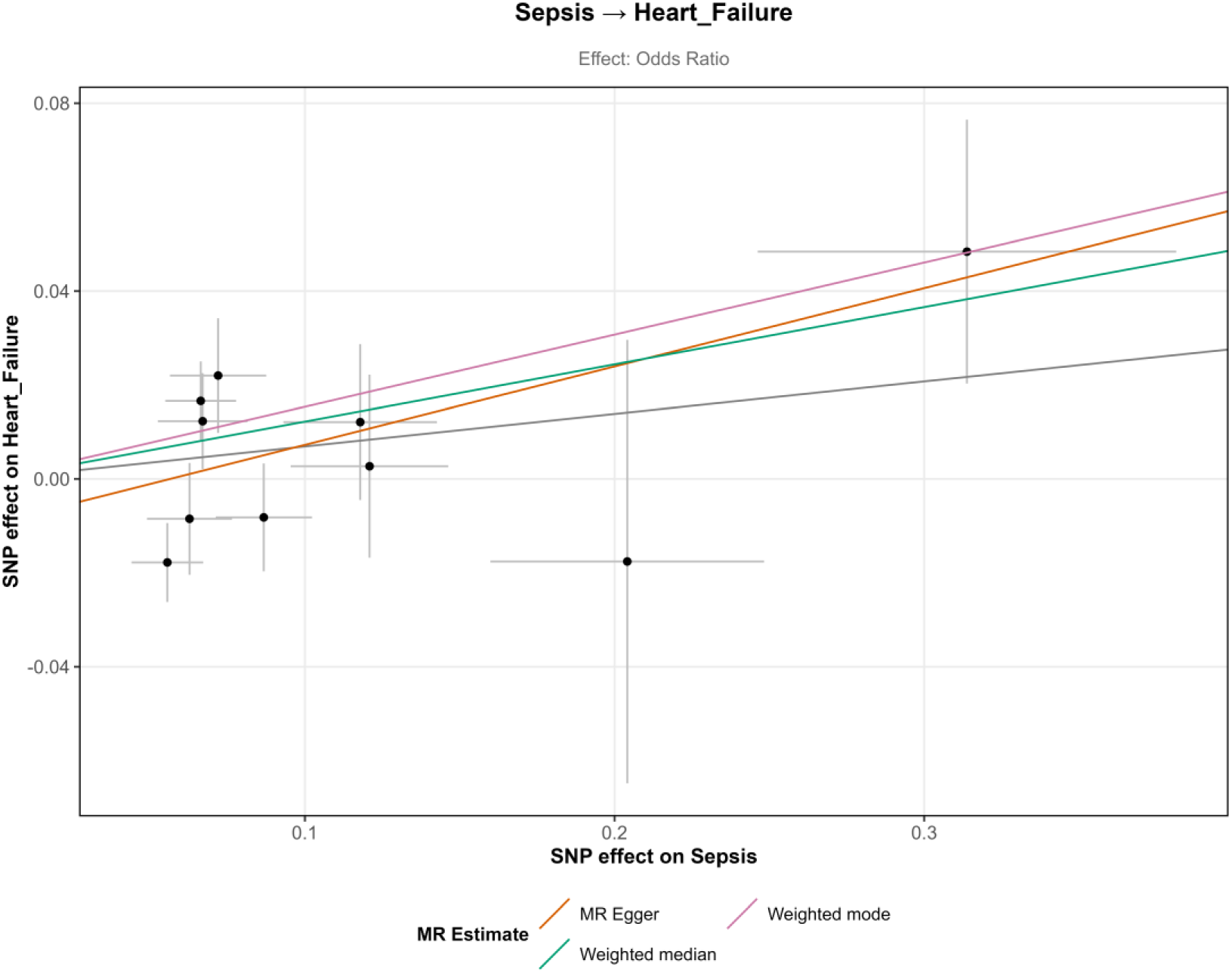
Scatter plot for the causal effect of sepsis on heart failure.

**Figure S4.**
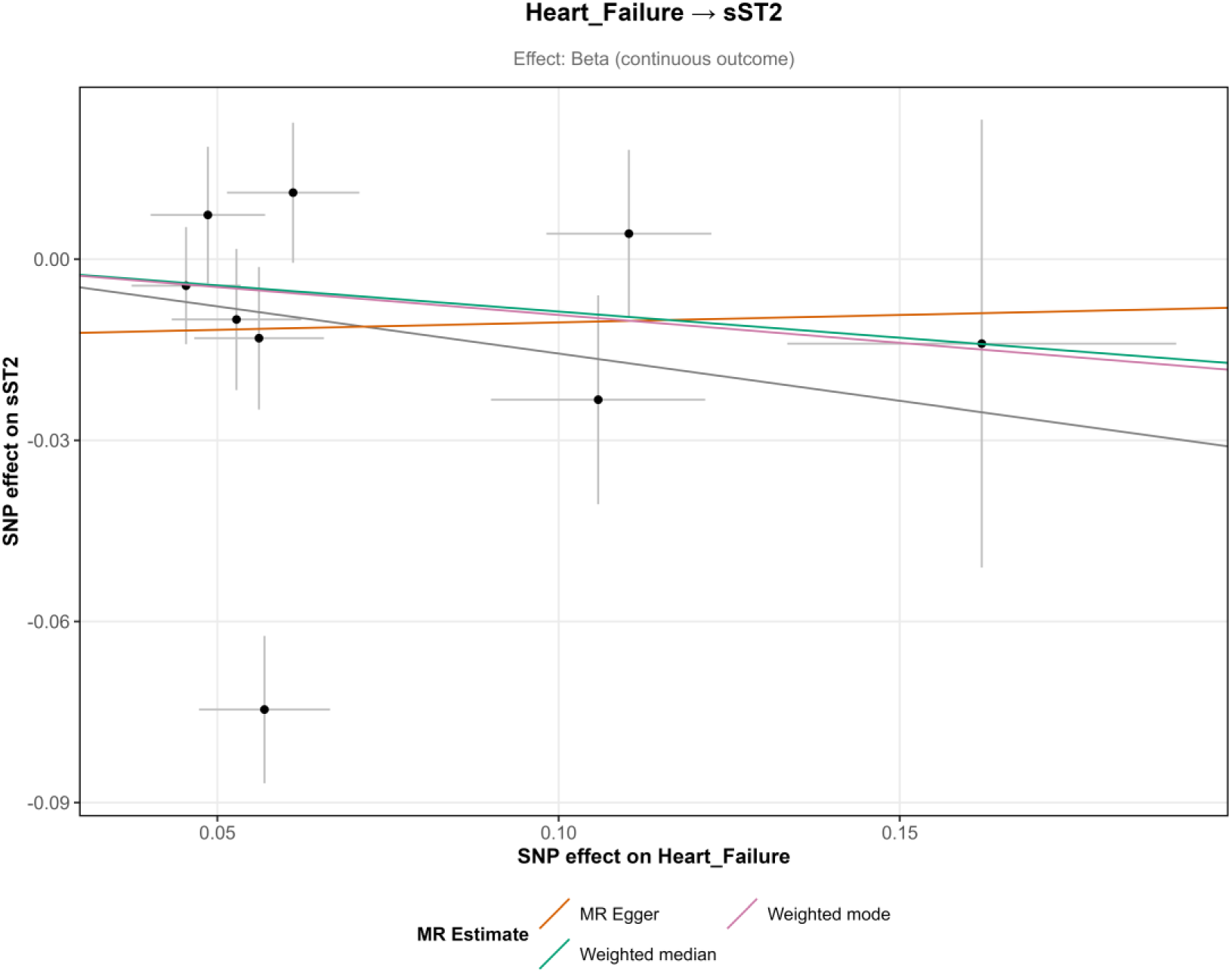
Scatter plot for the causal effect of heart failure on sST2 levels.

**Figure S5.**
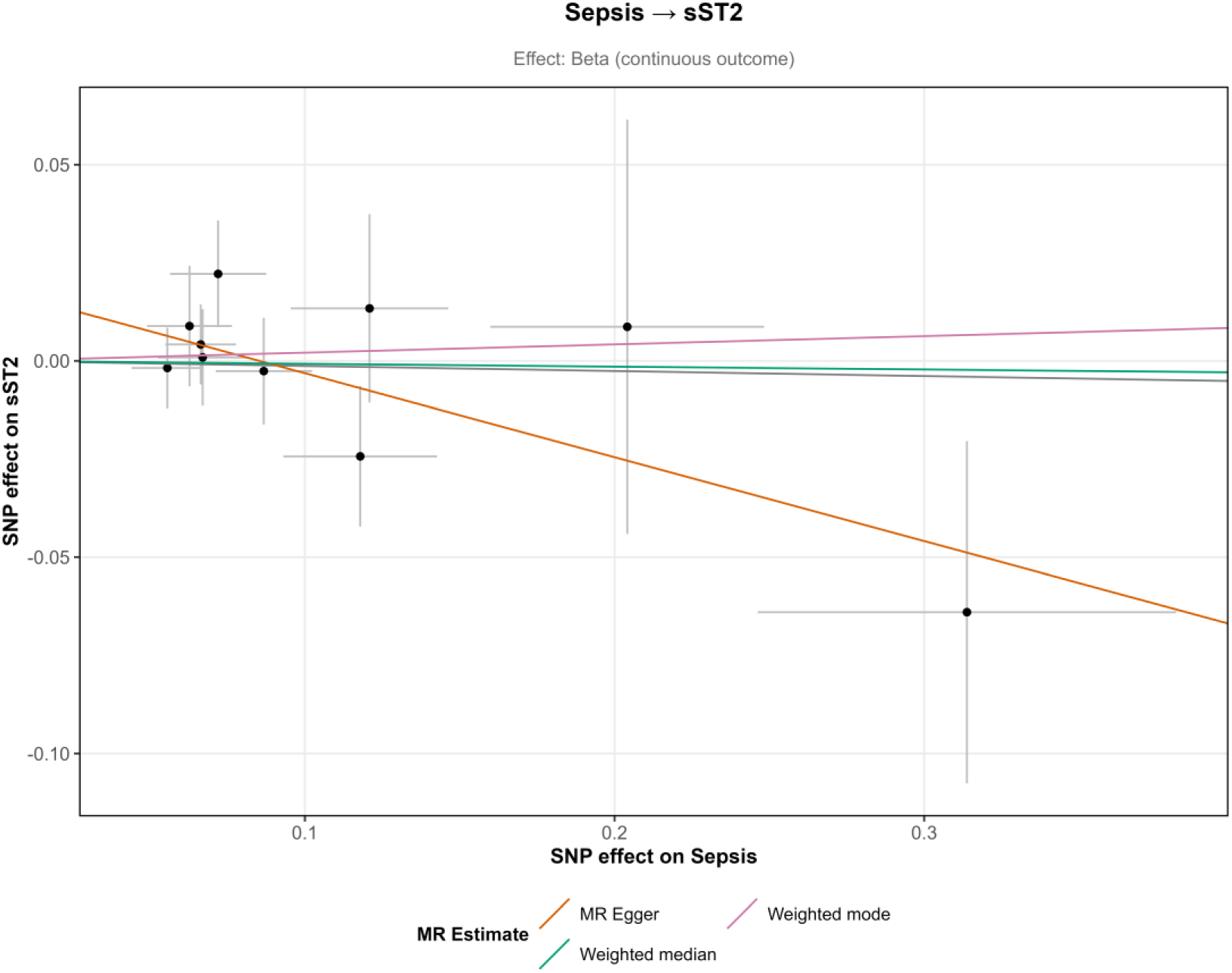
Scatter plot for the causal effect of sepsis on sST2 levels.

**Table S1.**
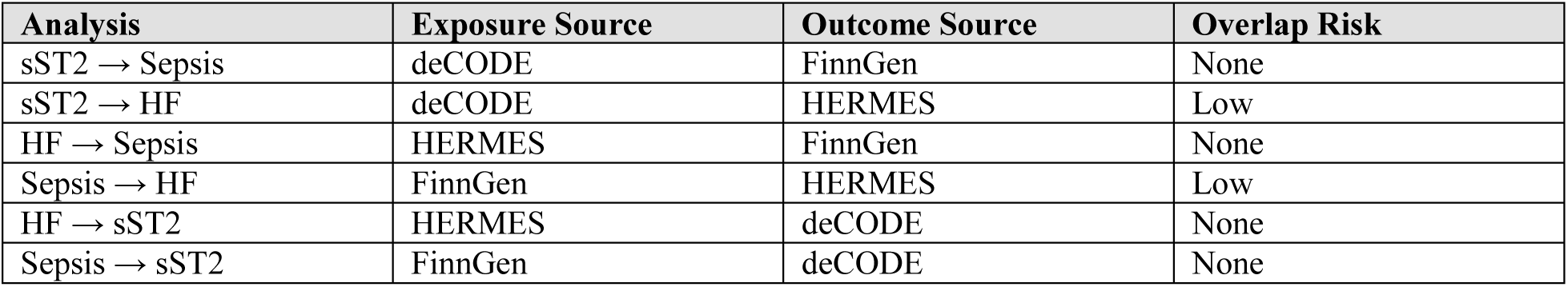
Assessment of Sample Overlap Between GWAS Datasets

| Analysis | Exposure Source | Outcome Source | Overlap Risk |
| --- | --- | --- | --- |
| sST2 → Sepsis | deCODE | FinnGen | None |
| sST2 → HF | deCODE | HERMES | Low |
| HF → Sepsis | HERMES | FinnGen | None |
| Sepsis → HF | FinnGen | HERMES | Low |
| HF → sST2 | HERMES | deCODE | None |
| Sepsis → sST2 | FinnGen | deCODE | None |

**Table S2.** Detailed Characteristics of cis-SNP Genetic Instruments for sST2.

| SNP | Chr | Position | Beta_Exp | SE_Exp | F-stat | OR | P-value |
| --- | --- | --- | --- | --- | --- | --- | --- |
| rs10204837 | 2 | 102977730 | 0.263 | 0.010 | 706.8 | 1.043 | 0.330 |
| rs1115282 | 2 | 102890970 | 0.337 | 0.010 | 1231.6 | 1.027 | 0.430 |

*cis*-SNPs are located within $\pm 500\text{kb}$ of the *IL1RL1* gene (chr2: 102,000,000-103,500,000 bp). Beta\_Exp, effect size on sST2; SE\_Exp, standard error; F-stat, F-statistic for instrument strength; OR, Wald ratio odds ratio for sepsis outcome.

## Notes

### Competing Interest Statement

The authors have declared no competing interest.

### Clinical Trial

Not applicable. This is a Mendelian Randomization study using secondary analysis of publicly available summary-level data and is not a prospective interventional study or clinical trial

### Author Declarations

This study is a secondary analysis of publicly available summarized genetic data. Ethical approval and participant consent were obtained by the original investigators of the respective primary GWAS consortia (deCODE Genetics, HERMES, and FinnGen)

